# Improving maternal, newborn, and child health outcomes through a community-based women’s health education program (*Chamas for Change*): a cluster randomized controlled trial

**DOI:** 10.1101/2020.06.28.20141663

**Authors:** Lauren Y. Maldonado, Jeffrey Bone, Michael L. Scanlon, Gertrude Anusu, Sheilah Chelagat, Anjellah Jumah, Justus E. Ikemeri, Julia J. Songok, Astrid Christoffersen-Deb, Laura J. Ruhl

## Abstract

**Introduction:** Community-based women’s health education groups may improve maternal, newborn, and child health (MNCH); however, evidence from sub-Saharan Africa is lacking. *Chamas for Change (Chamas)* is a community health volunteer (CHV)-led health education program for pregnant and postpartum women in western Kenya. We evaluated *Chamas*’ effect on facility-based deliveries and other MNCH outcomes.

**Methods:** We conducted a cluster randomized controlled trial involving 74 communities in Trans Nzoia County. We included pregnant women who presented to health facilities for their first antenatal care visits by 32 weeks gestation. We randomized community clusters 1:1 without stratification or matching; we masked data collectors, investigators, and analysts to allocation. Intervention clusters were invited to bimonthly, group-based, CHV-led health lessons (*Chamas*); control clusters had monthly CHV home-visits (standard of care). The primary outcome was facility-based delivery at 12-months follow-up. We conducted an intention-to-treat approach with multilevel logistic regression models using individual-level data. We prospectively registered this trial with ClinicalTrials.gov (NCT03187873).

**Results:** Between November 27, 2017 and March 8, 2018, we enrolled 1920 participants from 37 intervention and 37 control clusters. A total of 1550 (80.7%) participants completed the study with 822 (82.5%) and 728 (78.8%) in the intervention and control arms, respectively. Facility-based deliveries improved in the intervention arm (80.9% vs 73.0%; Risk Difference (RD) 7.4%, 95% CI 3.0-12.5, OR=1.58, 95% CI 0.97-2.55, p=0.057). *Chamas* participants also demonstrated higher rates of 48-hour postpartum visits (RD 15.3%, 95% CI 12.0-19.6), exclusive breastfeeding (RD 11.9%, 95% CI 7.2-16.9), contraceptive adoption (RD 7.2%, 95% CI 2.6-12.9), and infant immunization completion (RD 15.6%, 95% CI 11.5-20.9).

**Conclusion:** *Chamas* participation was associated with significantly improved MNCH outcomes compared with the standard of care. This trial contributes robust data from sub-Saharan Africa to support community-based, women’s health education groups for MNCH in resource-limited settings.

**KEY QUESTIONS:** *What is already known?:*

- Globally, maternal and infant deaths have declined over the last three decades; however, low and middle-income countries (LMICs), including Kenya, still disproportionately incur the highest morbidity and mortality.
- The World Health Organization recommends leveraging lay health workers (LHWs), including community health volunteers (CHVs), to promote maternal, newborn, and child health (MNCH) in resource-limited settings.
- Prior research suggests coupling strategies that promote community-based approaches (i.e. integrating LHWs) and women’s health education and support groups during pregnancy and postpartum may improve MNCH; however, robust evidence from sub-Saharan Africa is lacking.

*What are the new findings?:*

- Using a cluster randomized controlled trial design, we found that participation in *Chamas for Change (Chamas)* – a group-based women’s health education program led by CHVs – was associated with significantly improved MNCH outcomes, including facility-based deliveries, compared with the standard of care (i.e. monthly home-visits) in rural Kenya.
- This trial also demonstrated significant associations between program participation and receiving 48 hour postpartum home-visits, breastfeeding exclusively, adopting a contraceptive method postpartum, and immunizing infants fully by 12 months of life as compared to the standard of care.
- These findings support pilot data from a preceding evaluation of the *Chamas* program as well as the current literature on community-based interventions delivered by LHWs to promote MNCH in other resource-limited settings.

*What do the new findings imply?:*

- Effective community-based strategies that build upon existing infrastructure to promote MNCH are needed to continue to improve the health and well-being of women and infants in rural sub-Saharan Africa and other LMICs.
- *Chamas* offers an innovative approach to improve MNCH in resource-limited settings with significant health policy implications; collective evidence from this trial and preceding studies support community-based women’s health education groups as an effective strategy for improving uptake of facility-based deliveries and other life-saving MNCH practices.

## Introduction

Globally, maternal and infant deaths have declined over the last three decades; however, low and middle-income countries (LMICs) still disproportionately incur the highest morbidity and mortality. Kenya’s maternal mortality ratio (MMR) and infant mortality rate (IMR) remain among the highest in the world at 342 per 100,000 live births and 31 per 1,000 live births, respectively.^1,2^ Fragile health systems, poor access to high quality and specialized care, low health literacy rates, gender-based inequities, and generational poverty contribute to this disparity.^3-5^ Effective solutions that build upon infrastructure to promote the health and well-being of women and infants are needed to continue to improve maternal, newborn, and child health (MNCH) outcomes in resource-limited settings.

Mobilizing community health volunteers (CHVs) to promote MNCH offers a promising strategy to reduce health inequities.^6-8^ In 2006, the Republic of Kenya Ministry of Health (MOH)’s “Kenya Essential Package for Health” delineated a comprehensive strategy to improve the health of households and communities, commonly known as the “Community Health Strategy” (CHS).^9^ Under the current CHS, CHVs are expected to perform monthly home-visits for all pregnant women during pregnancy and postpartum.^10^ Despite these efforts, the practice of key MNCH interventions associated with demonstrated reductions in mortality and morbidity are well-below projected targets to substantively reduce the MMR and IMR.^1^ These gaps are pronounced across socio-economic and geographic strata with women in poorer, rural communities experiencing significantly worse outcomes than those in wealthier, urban centers.

The World Health Organization (WHO) recommends integrating lay health workers, including CHVs, to promote MNCH interventions.^11^ Coupling this strategy with the delivery of group-based women’s health education may improve MNCH; however, evidence from sub-Saharan Africa is lacking.^12^ In 2012, the Academic Model Providing Access to Healthcare (AMPATH) - a long-standing partnership between the Kenyan Ministry of Health (MOH), Moi University, Moi Teaching and Referral Hospital, and North American universities - launched *Chamas for Change* (*Chamas*) to address this unmet need. *Chamas* is a CHV-led health education program that supports women during pregnancy and for the first 1,000 days of their child’s life. The program hybridizes best practices from resource-limited settings globally to offer a community-based, multi-pronged strategy for improving MNCH. This strategy focuses on providing health education, a peer-supportive environment, and opportunities to access financial capital to promote MNCH while simultaneously addressing inequities that perpetuate poor outcomes.

A pilot study evaluating the first year of *Chamas* demonstrated significant associations between participation and the likelihood of practicing positive MNCH behaviors.^13^ To determine whether first-year *Chamas* participation could improve MNCH outcomes, including facility-based delivery, we conducted a cluster randomized controlled trial in rural western Kenya.

## Methods

### Study design

We conducted a two-arm cluster randomized controlled trial in 74 communities across four sub-counties (Cherangany, Kwanza, Kiminini, Saboti) in Trans Nzoia County, Western Province, Kenya (**Figure 1**). Cluster randomization was used to avoid potential contamination of intervention activities between neighboring villages. We prospectively registered this trial with ClinicalTrials.gov (NCT03187873). We received ethics approvals from the Institutional Research Ethics Committee at Moi University and Moi Teaching and Referral Hospital (IREC/2018/269) and Institutional Review Board at Indiana University (1905296355). We obtained written informed consent from all participants prior to data collection.

**Figure 1.**
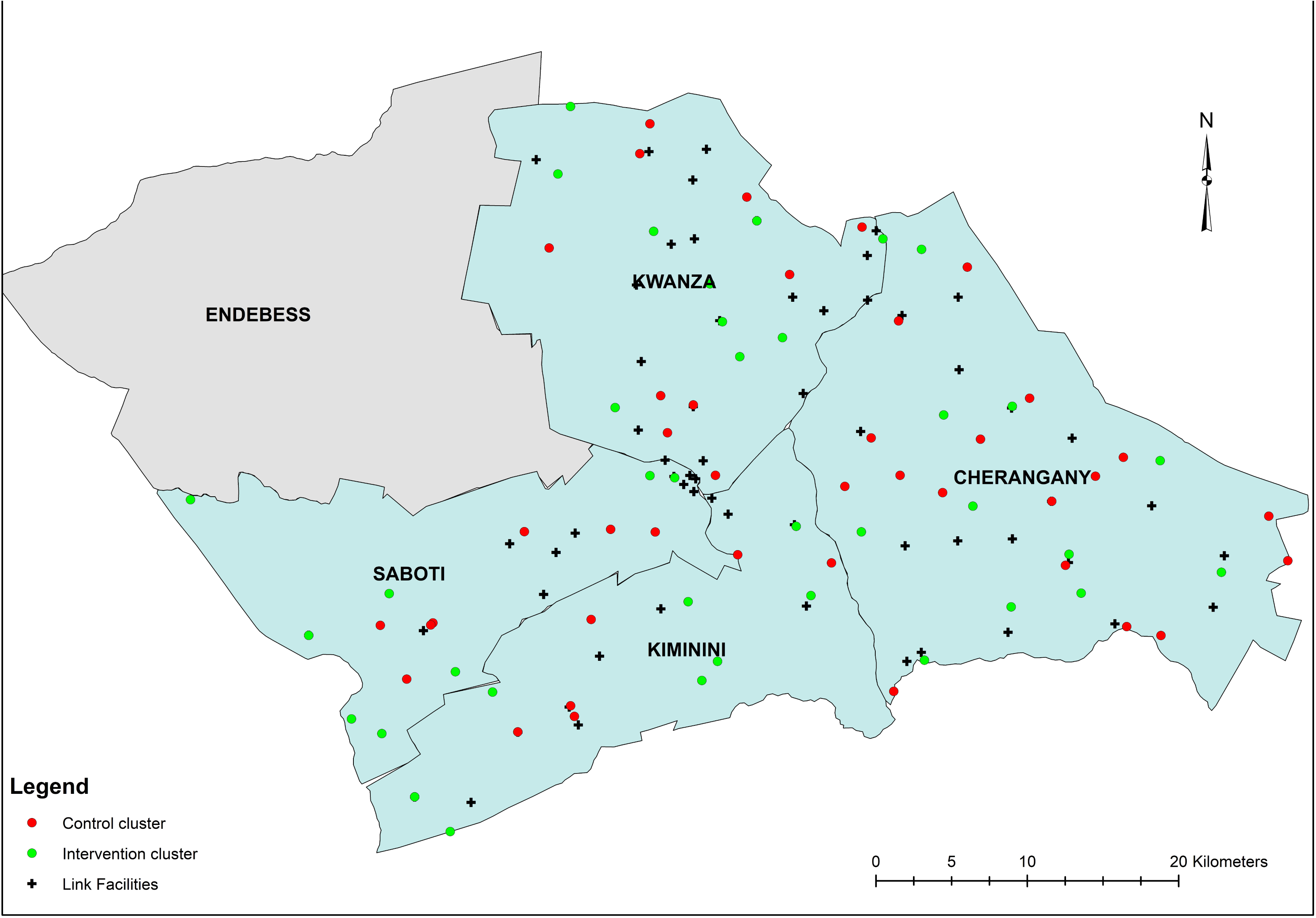
Cluster map.

### Participants

We identified 77 community health units (CUs) among 163 total CUs to serve as potential clusters. CUs are geographically defined health service delivery areas for populations of 5,000 people supervised by Community Health Extension Workers (CHEWs) and CHVs. These CUs were specifically chosen as their CHVs received formal CHS training from AMPATH. We recruited participants from 60 public and private health facilities that provide care for these CUs. Pregnant women who were less than or equal to 32 weeks gestation, presenting for their first antenatal care (ANC) visits, and residing in one of the 77 CUs were eligible. We selected a gestational age cut-off of 32 weeks as the majority (96.0%) of Kenyan women who seek antenatal care at any point during pregnancy present for at least one ANC visit by this time.^1^ Due to slow recruitment resultant of preceding health worker strikes in Trans Nzoia, we increased our original gestational cut-off from 28 to 32 weeks.

### Randomization and masking

We randomized CUs 1:1 to intervention (e.g., *Chamas* program) or standard of care (e.g., monthly CHV home-visits). The trial data manager used a simple random allocation sequence generated by *PASS 11 (version 11*.*0*.*10)* to designate cluster assignment. Non-study CUs served as buffer zones between intervention and control clusters to avoid contamination. There was no stratification or matching. We masked data collectors, investigators, and analysts to cluster allocation throughout the trial; however, both arms were identifiable to participants and CHVs by design.

### Procedures

Data collectors assessed women for eligibility at their first ANC visit. Women deemed eligible and willing to participate provided consent to be contacted for enrollment. The trial data manager generated lists of participants organized by residential CUs. These lists were subsequently distributed to CHVs who were tasked with finding women in their communities and enrolling them. Data collectors accompanied CHVs during this process and obtained baseline data at enrollment. One week following the end of the enrollment period, the data manager randomized all CUs to intervention and control arms. Three weeks later, CHVs began facilitating *Chamas* or delivering home-visits across clusters.

Intervention clusters participated in the *Chamas* program. Program details are published elsewhere.^13^ Briefly, *Chamas* is a group-based, CHV-led health education program that supports women during pregnancy and for the first 1,000 days of their child’s life. Participants attend 60-90 minute sessions twice a month, which include discussions on health and social topics relevant to antenatal, postpartum, and early childhood experiences. CHVs use an illustrated flip-chart with evidence-based, structured discussion guides to facilitate lessons. Groups are typically comprised of 15-20 women, two CHV facilitators, and two mentor mothers (e.g. post-menopausal women who have completed child-rearing). The first year of the curriculum promotes behaviors associated with demonstrated reductions in maternal and infant morbidity and mortality, including but not limited to: attending ANC visits, delivering in health facilities, and exclusively breastfeeding. These lessons purposefully mirror health topics CHVs are expected to promote during home-visits under the CHS. Following each lesson, women are invited to participate in an optional table-banking program called *Group Integrated Savings for Health and Empowerment (GISHE). GISHE* participation is optional so as not to deter women without financial means to contribute to group savings from joining *Chamas*. Women are encouraged to use savings generated by *GISHE* to finance health interventions (e.g., enroll in health insurance, pay for transportation to health facilities), invest in early childhood education, and/or start small businesses.

Strategies to ensure fidelity of *Chamas* included: using standardized intervention materials (i.e. printed curriculum flipcharts), hosting structured CHV training sessions preceding the trial, offering monthly supervision by study staff, and designating at least two trained CHVs to every group to avoid potential disruptions due to illnesses or job transfers. In addition to attending the four-day MNCH refresher training, CHVs facilitating *Chamas* also received a formal two-day orientation to the program and were trained in group facilitation techniques. We provided scheduled support sessions for CHV facilitators throughout the trial (during months 1-3, 6, 9, and 12), which provided opportunities for feedback and communal trouble-shooting to enhance program delivery.

Control clusters had monthly CHV home-visits during pregnancy and postpartum, as recommended by the Kenyan CHS standard of care.^10^ During monthly visits, CHVs collect basic health information, identify antenatal and early postpartum danger signs, refer individuals to care (if indicated), and aid in infant growth monitoring. CHVs are also expected to encourage women to adopt the same key health behaviors promoted in *Chamas*. CHVs working within control clusters received oversight and supervision from CHEWs, as structured by the CHS. We did not provide incentives (monetary or other) to CHVs, CHEWs or participants in either study arm at any point during the trial.

### Outcomes

We measured outcomes at the individual-level. We selected facility-based delivery as our primary outcome because of the significant association between institutional delivery and reductions in maternal and infant morbidity and mortality.^14-16^ Secondary outcomes included: attending at least four ANC visits, receiving a 48-hour postpartum home-visit, exclusively breastfeeding for six months, adopting a modern contraceptive method, immunizing infants with the oral polio (OPV 0) vaccine within two weeks postpartum, immunizing infants with the measles vaccine (Measles I) by 12 months of age, and completing the infant immunization series per WHO and Republic of Kenya MOH standards by 12 months of age.^17-19^

Data collectors travelled to participant homes to collect end-line data 12 months following the initiation of *Chamas* sessions and home-visits. Outcome measures were self-reported with the exception of infant immunizations, which were extracted from standard MOH Maternal Child Health Booklets kept by mothers. All data were recorded using electronic, standardized questionnaires. We classified participants as lost to follow-up after we made three attempts to establish contact over a two-week period. We conducted abbreviated phone surveys if participants relocated outside of Trans Nzoia County; these abbreviated questionnaires omitted questions on infant immunizations.

At enrollment, we collected baseline participant socio-demographic (age, marital status, maternal education, occupation, poverty probability index scores, insurance status) and reproductive health (previous pregnancy and related outcomes) data. We used the Kenya 2015 *Poverty Probability Index (PPI)* questionnaire and national poverty line scorecard to estimate participants’ poverty likelihood at baseline.^20^ We recorded attendance at each *Chamas* session to track individual program participation. A Data and Safety Monitoring Board recorded and investigated adverse events including CHV-reported participant mortalities as well as the cause of death (if known).

### Statistical analysis

We estimated sample size using methods described by *Rutterford et al*. for a proposed mixed effects regression analysis^21^ using derived baseline estimates.^1,13^ Assuming a mean cluster size of 20 individuals, 77 clusters (equally allocated between arms), intra-cluster correlation coefficient (ICC) of 0.44 (based on pilot data^13^), and 20% attrition, we calculated a total of 1,280 individuals would be needed to detect a 4.7% difference on the risk difference scale with 80% power at a (two-tailed) significance level of 0.05. To determine our recruitment timeline, we assumed 6.3% of all women of reproductive age would be pregnant at any given time (or roughly 50 women per CU annually).^1^ We determined an enrollment period of roughly 3-4 months adequate to recruit our estimated sample size.

Our primary analyses were intention-to-treat (ITT) and included all participants from randomized clusters who provided baseline and 12-month follow-up data, regardless of the level of participation in *Chamas*. We summarized all demographic and reproductive health history information between arms with means and standard deviations for continuous variables and counts and percentages for categorical variables. We analyzed the primary outcome with multilevel logistic regression with a random intercept for cluster, and effects are presented as risk differences with 95% bootstrap confidence intervals and odds ratios with 95% Wald-type confidence intervals and p-values. We also report the intra-cluster correlation coefficient (ICC). We analyzed secondary outcomes similarly.

For both primary and secondary outcomes, we conducted several sensitivity analyses. First, to assess the impact of missing outcomes due to lost to follow-up, we used multiple imputation with 10 datasets with the ‘jomo’ algorithm to account for the multilevel structure of the data; results were then combined using Rubin’s rules.^22,23^ Second, to assess the possible impact of differences in factors known to be associated with care seeking behaviors between arms, we adjusted our primary models for: PPI score, marital status, null parity and health insurance at time of delivery. A third sensitivity analyses combined adjustment and imputation. Lastly, we performed a sensitivity analysis restricting our intervention sample to women who attended at least one *Chamas* session during the trial period.

We assessed the effect of *Chamas* participation on infant vaccination outcomes similarly, but given the large amount of missing data, no sensitivity analyses with imputation were conducted. Adjusted models for vaccination only included maternal education, PPI, and insurance at delivery as additional covariates have not been shown to be associated with vaccination rates. Further, since vaccination data was missing in approximately 40% of the sample, we were concerned about selection bias in those reporting the outcome. To account for this, we carried out an additional sensitivity analysis to indicate the amount of unmeasured confounding between trial arm and vaccination that would be needed to explain away the observed differences.^24^

There were no interim analyses. We developed, finalized, and signed a statistical analysis plan prior to beginning data analysis (**Supplement, Statistical Analysis Plan**). Statistical significance was set at 0.05 and all analyses were conducted using R statistical software (version 3.5.3).^25^

### Patient and public involvement

To honor active community involvement, we sought and incorporated feedback from a multidisciplinary study advisory committee, including direct beneficiaries (i.e. participating women, CHVs) and key stakeholders (i.e. local community leaders, Kenyan MOH representatives, research advisors), in the initial design and conception of this trial. We designed our questionnaires, data instruments, and intervention activities based on qualitative feedback provided by program participants during pilot studies of the *Chamas* program. These qualitative questionnaires attempted to capture participant perceptions of the strengths and weaknesses of the program, as well as priority areas for continued improvement. Prior to initiating trial activities, we invited CHVs, CHEWs, health facility managers, sub-county MOH representatives, and community leaders to stakeholder meetings to explain the study’s purpose and procedures, as well as to facilitate understanding of our trial objectives among leadership at the county, sub-county, and community levels. Following these meetings, we asked community leaders for permission to begin enrolling participants. All CHVs who agreed to participate also attended a four-day refresher training on their roles and expectations in promoting MNCH under the Kenyan CHS. We discussed the trial’s risks and benefits with all participants before enrollment, including demands on individual time due to program participation and data collection. We obtained written informed consent from all participants prior to data collection. At the trial’s conclusion, we verbally disseminated our preliminary findings to the program’s direct beneficiaries and key stakeholders. We plan to additionally distribute printed summaries of key findings following publication of this trial.

### Role of the funding source

The funders had no role in the research design, collection, analysis, or interpretation of data, writing this report, or the decision to submit this manuscript for publication. The corresponding author had full access to all data in the study as well as final responsibility for the decision to submit this manuscript for publication.

## Results

Details of our enrollment and inclusion procedures are summarized in **Figure 2**. Between November 27, 2017 and March 8, 2018, we assessed 4235 women for eligibility; 2923 women from 74 clusters met criteria and agreed to be contacted. CHVs successfully contacted and enrolled 1920 eligible women from 74 community clusters (996 participants in 37 intervention and 924 in 37 control clusters). We collected follow-up data on all clusters between April 7 and July 3, 2019. A total of 1550 (80.7%) participants completed the study at 12-month follow-up: we included 822 in the intervention (82.5%) and 728 in the control (78.8%) arms for analysis. Among 822 intervention participants who completed the study, 599 (72.9%) attended at least one *Chamas* session. Among those who attended, mean attendance was 12 (SD 7.8) of 24 total sessions and 48.9% participated in *GISHE*. Participants who were lost to follow-up were similar in number across study groups and attrition was not associated with sociodemographic or reproductive health characteristics.

**Figure 2.**
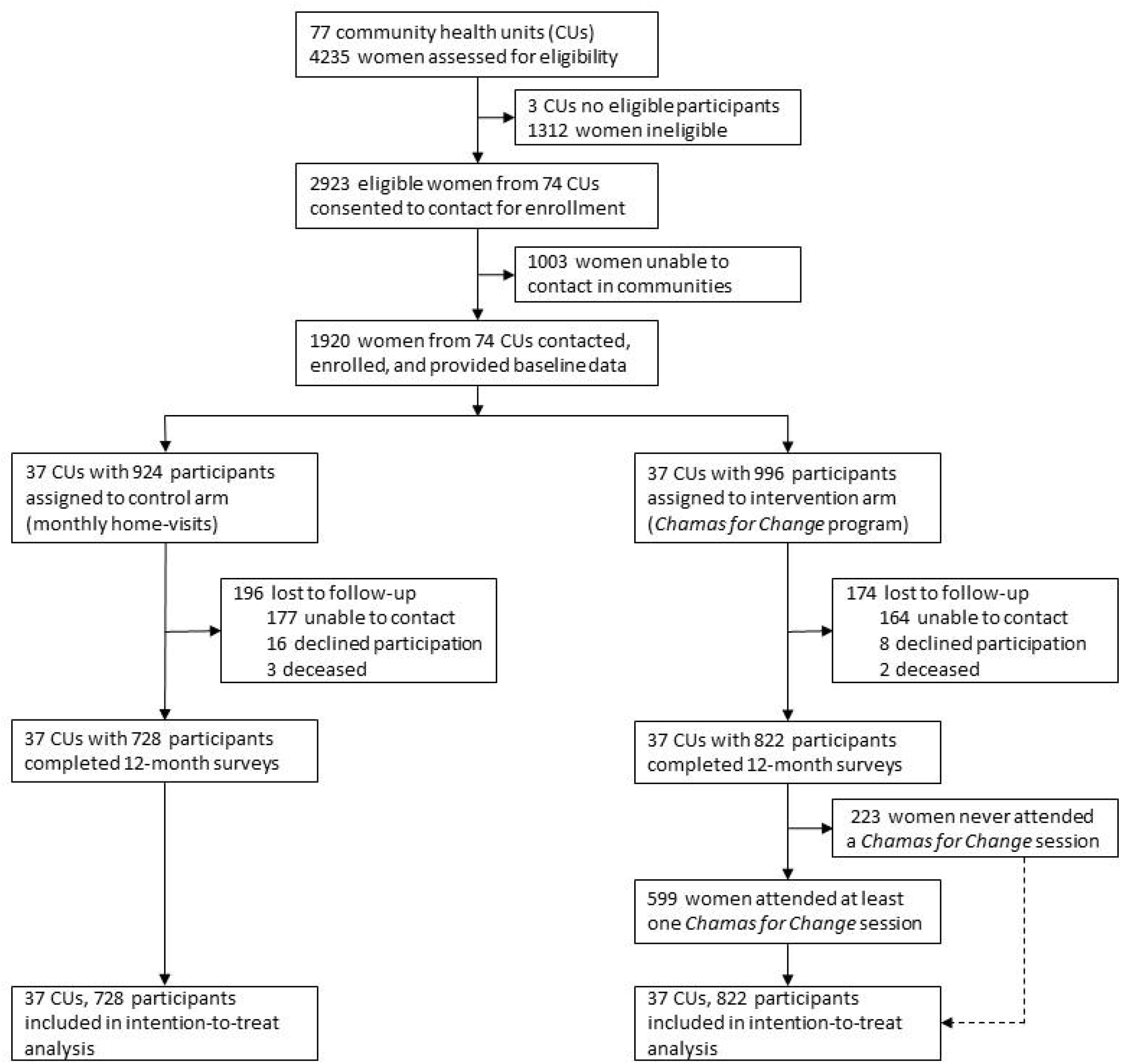
Trial Profile.

Participants who completed the study (n=1550) were similar in baseline characteristics (**Table 1**). Most participants were married, unemployed, completed primary school, possessed health insurance at the time of delivery, and carried a previous pregnancy. The mean PPI score for our study population was 55.13 (SD 20.11); PPI scores differed across study arms with higher values among control compared to intervention participants at baseline. Cluster-level demographics were also well-balanced. Across all clusters, CHVs possessed a mean 11.69 (SD 6.32) years of experience. Lastly, in the intervention arm, we noted geographic differences among women who attended *Chamas* and those who never attended (**Table S2**).

**Table 1.**
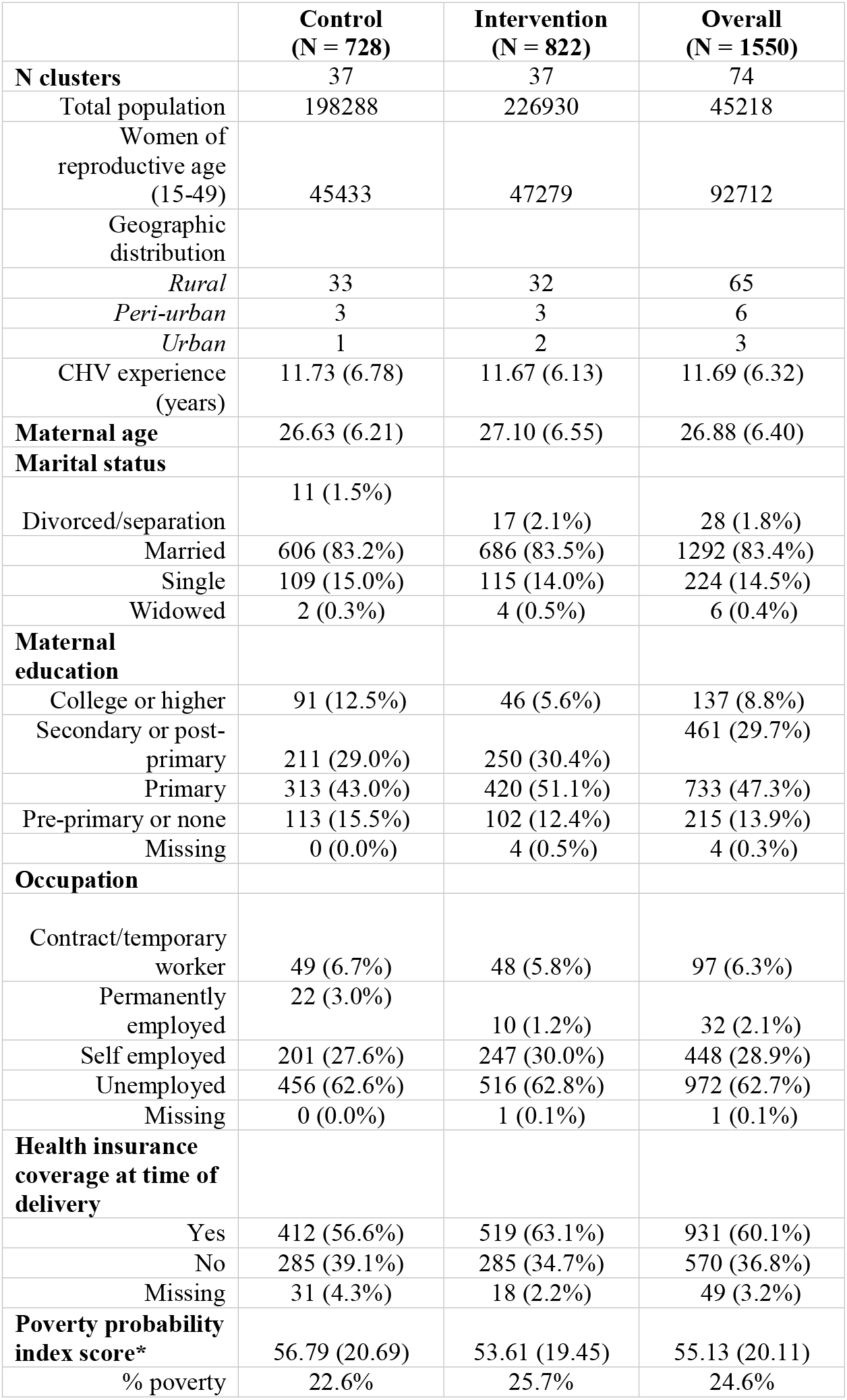
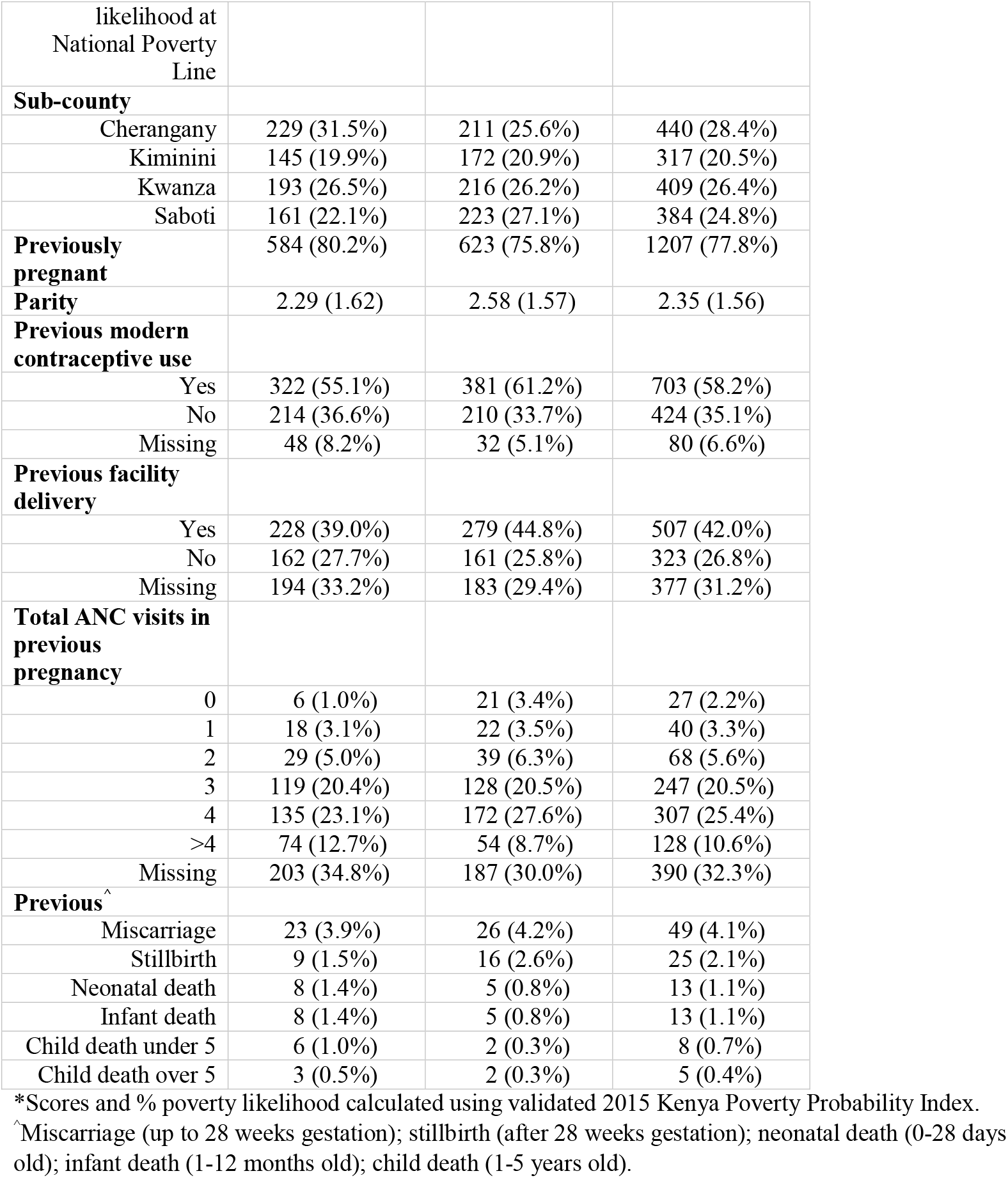
Baseline characteristics for intention-to-treat population (74 clusters, n=1550)

Primary and secondary outcomes are summarized in **Table 2**. The overall proportion of health facility delivery was higher among intervention (80.9%, 653 participants) than control participants (73.0%, 514 participants). Among women who did not deliver in a health facility (n=383), the most commonly cited reasons across cohorts included: preference to deliver at home or with a traditional birth attendant (32.1%), structural challenges associated with reaching a health facility (e.g. too far, poor road conditions) (32.1%), and medical emergencies (e.g. abrupt labor with not enough time to travel) (11.5%). In unadjusted models, we estimated a 7.4% (95% CI 3.0-12.5) improvement in facility-based deliveries (OR=1.58, 95% CI 0.97-2.55, p=0.057). Following adjustment and adjustment with imputation, this improvement was slightly attenuated to 6.4% (95% CI 2.0-10.4) and 7.1% (95% CI 3.0-11.4), respectively (aOR1=1.59 95% CI 1.02-2.47, p=0.042; aOR2=1.62 95% CI 1.06-2.49, p=0.004) (**Table S3**). Further, a sensitivity analysis restricting the intervention sample to women who attended *Chamas* at least once attenuated improvement in facility-based delivery by 5.2% (95% CI 1.5-9.5) (OR=1.43 95% CI 0.92-2.24, p=0.11) (**Table S3**). We observed a relatively large amount of cluster heterogeneity as indicated by an ICC of 0.18 (**Figure 3**).

**Table 2.** Primary and secondary outcomes: facility-based delivery, care seeking and vaccination.

|  | Control | Intervention | Risk difference<br>(95%CI) | Odds ratio<br>(95%CI) | p-value* |
| --- | --- | --- | --- | --- | --- |
| Facility-based delivery | 514 (73.0%) | 653 (80.9%) | 7.4% (3.0%, 12.5%) | 1.58 (0.969, 2.55) | 0.057 |
| Adequate ANC care | 507 (69.6%) | 587 (71.4%) | 3.2% (-1.5%, 7.7%) | 1.18 (0.82,1.68) | 0.375 |
| Postnatal CHV visit | 97 (13.6%) | 241 (30.1%) | 15.3% (12.0%, 19.6%) | 3.22 (1.50,6.93) | 0.003 |
| Exclusive breast feeding for 6 months | 383 (56.7%) | 521 (67.2%) | 11.9% (7.2%, 16.9%) | 1.77 (1.12,2.80) | 0.014 |
| Contraceptive use | 472 (65.5%) | 581 (71.8%) | 7.2% (2.6%, 12.9%) | 1.41 (1.03,1.93) | 0.034 |
| Long-acting reversible contraceptive use | 242 (51.3%) | 326 (56.1%) | 7.1% (0.9%, 13.3%) | 1.34 (0.95,1.91) | 0.099 |

\*p-value is for odds ratio from mixed effect logistic regression.
\*\*Denominators are based on number of women reporting the particular outcome. See Table S1 for details.

**Figure 3.**
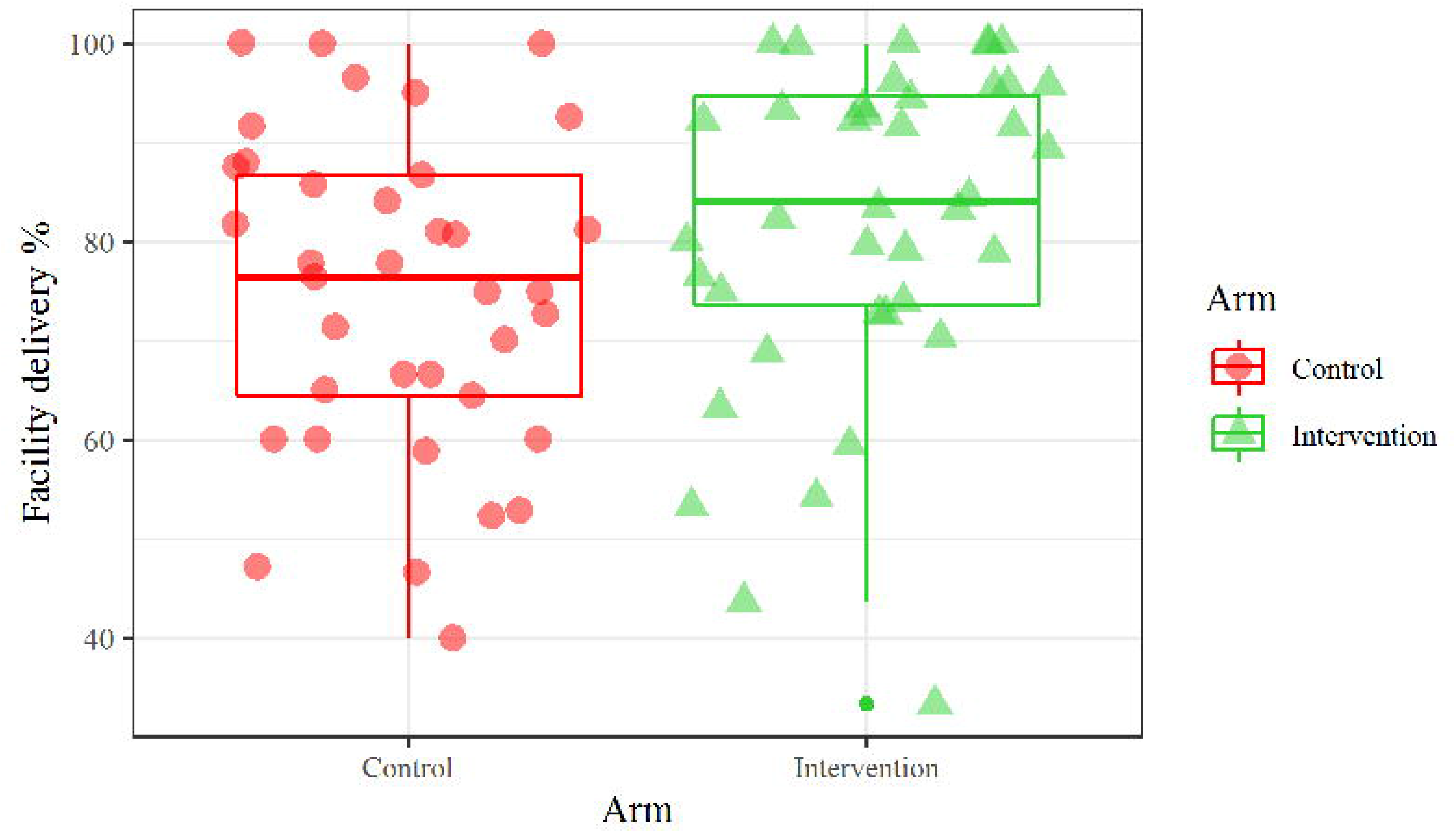
Cluster outcome rates.

We examined the effect of *Chamas* participation on secondary MNCH outcomes associated with demonstrated reductions in maternal and infant morbidity and mortality. Women in *Chamas* clusters improved in 48-hour postpartum visits (15.3%, 95% CI 12.0-19.6), exclusive breastfeeding (11.9%, 95% CI 7.2-16.9), and contraceptive adoption (7.2%, 95% CI 2.6-12.9) compared with controls (**Table 2**). Though not statistically significant, the risk differences in achieving adequate ANC and adopting a long-acting method of contraception (i.e. intrauterine device or implant) were also greater among *Chamas* participants. Restricting our intervention sample to women who attended *Chamas* at least once accentuated improvements in 48-hour postpartum visits (19.6%, 95% CI 14.4-25.0) and exclusive breastfeeding (13.6%, 95% CI 7.8-19.8); conversely, we observed an attenuated effect with this restriction on contraceptive adoption (5.7%, 95% CI 0.7-11.1) (**Table S3**). Other sensitivity analyses did not meaningfully change results (**Table S3**).

We additionally assessed infant immunization outcomes among live infants at follow-up. Infants born to women in *Chamas* demonstrated significant improvements in receiving the Measles I vaccine by 12 months of age (13.2%, 95% CI 9.1-18.4) and completing the recommended infant immunization series per WHO (15.6%, 95% CI 11.5-20.9) and Republic of Kenya MOH (15.1%, 95% CI 10.4-20.3) guidelines (**Table 3**). These results were unchanged after adjusting for covariates (**Table S4**). We estimated an unmeasured confounder (due to selection bias in those that reported the outcome) associated with both increased rate of vaccination and enrollment in intervention trial arm (compared to control) by 30% would be required to explain away these observed significant differences.

**Table 3.** Infant immunization outcomes.

|  | Control | Intervention | Risk difference<br>(95%CI) | Odds ratio<br>(95%CI) | p-value* |
| --- | --- | --- | --- | --- | --- |
| Infants who received OPV 0 within 2 weeks of birth | 341 (64.6%) | 361 (66.0%) | 1.7% (-3.6%, 8.4%) | 1.08 (0.77,1.51) | 0.663 |
| Infants who received Measles I by 12 months of age | 328 (74.0%) | 339 (87.6%) | 13.2% (9.1%, 18.4%) | 2.71 (1.45,5.04) | 0.002 |
| Fully-immunized infants ($\leq 12$ months) per WHO standards | 324 (73.6%) | 352 (88.9%) | 15.6% (11.5%, 20.9%) | 3.52 (1.74,7.12) | < 0.001 |
| Fully-immunized infants ($\leq 12$ months) per Republic of Kenya MOH standards | 320 (73.1%) | 348 (87.7%) | 15.1% (10.4%, 20.3%) | 3.16 (1.61,6.21) | < 0.001 |

\*p-value is for odds ratio from mixed effect logistic regression.
\*\*Denominators are based on number of women reporting the particular outcome. See Table S1 for details.

Maternal and infant mortality and morbidity outcomes are presented in **Table 4** with no significant differences between trial arms; however, the trial was not powered to detect differences in these relatively rare outcomes. Overall, we observed a protective effect of *Chamas* participation against maternal (−4.7%, 95% CI −9.4, 0.1) and infant (−3.9%, 95% CI −8.6, −0.3) morbidity. We recorded five participant mortalities during the trial (two in intervention and three in control). Three deaths were attributed to maternal causes of mortality, notably: one due to obstructed labor, one due to post-caesarian infection, and one due to eclampsia; the remaining two deaths were attributed to complications of cervical cancer. None of these mortalities were directly associated with trial participation. Across both trial arms, we recorded 43 perinatal deaths (i.e., deaths during the first week of life), 15 neonatal deaths, and 25 infant deaths.

**Table 4.** Maternal and infant mortality and morbidity outcomes.

|  | Control | Intervention | Risk difference<br>(95%CI) | Odds ratio<br>(95%CI) | p-value* |
| --- | --- | --- | --- | --- | --- |
| Maternal mortality | 3 (<0.1%) | 2 (<0.1%) | - | - | - |
| Maternal morbidity | 136 (18.7%) | 110 (13.4%) | -4.7% (-9.4%, 0.1%) | 0.68 (0.42, 1.10) | 0.118 |
| Miscarriage | 16 (2.2%) | 13 (1.6%) | -0.2% (-1.3%, 0.8%) | 0.85 (0.30, 2.38) | 0.751 |
| Stillbirth | 16 (2.2%) | 12 (1.5%) | -0.6% (-1.7, 0.3%) | 0.64 (0.27, 1.56) | 0.331 |
| Perinatal death | 22 (3.1%) | 21 (2.6%) | -0.5% (-1.9%, 0.8%) | 0.83 (0.42, 1.66) | 0.601 |
| Neonatal death | 6 (0.87%) | 9 (1.13%) | 0.2% (-0.6%, 0.7%) | 1.29 (0.39, 4.29) | 0.674 |
| Infant death | 13 (1.9%) | 12 (1.5%) | -0.2% (-1.3%, 0.8%) | 0.83 (0.27, 2.5) | 0.689 |
| Low birthweight | 118 (16.0%) | 157 (18.9%) | 1.9% (-1.6%, 5.6%) | 1.16 (0.70, 1.90) | 0.570 |
| Infant morbidity | 132 (18.68%) | 118 (16.74%) | -3.9% (-8.6%, -0.3%) | 0.76 (0.51, 1.15) | 0.194 |

\*Maternal morbidity defined as any health condition attributed to and/or aggravated by pregnancy and childbirth that has a negative impact on the woman's wellbeing, including the following complications: miscarriage (<28 weeks), stillbirth (>28 weeks), gestational diabetes, preeclampsia, eclampsia, postpartum infection, postpartum hemorrhage, or obstructed labor.
\*\*Infant morbidity defined as any health condition that affects mortality rate during the first-year of life including low birthweight (<2.5 kg), perinatal disorders (gestational diabetes, preeclampsia, eclampsia), infant immunization adherence, exclusive breastfeeding, and delivery-related complications (obstructed labor, neonatal resuscitation).
\*\*\*Perinatal deaths (first week of life), neonatal deaths (through 28<sup>th</sup> day of life), infant deaths (through first year of life).

## Discussion

In Kenya and other resource-limited settings, effective community-based strategies are increasingly needed to reduce maternal and infant deaths. Encouraging facility-based delivery is one well-known and highly effective strategy to achieve this goal.^26^ Despite government-led initiatives that support access to health services - such as the CHS and elimination of delivery-related fees at public facilities announced in 2013 - facility-based deliveries nationally (61.2%) and in our study area (58.3%) are still lower than needed to sufficiently reduce the MMR and IMR.^1,27^ Against this backdrop of under-utilized intrapartum services, we rigorously tested a community-based women’s health education program designed to improve facility-based deliveries and other MNCH practices. We evaluated outcomes in an intention-to-treat sample derived from a geographically diverse catchment area. These analyses produced findings that mimic real-world scenarios in which perfect program attendance is unlikely. We found that facility-based delivery and other key MNCH practices significantly improved in the intervention arm, providing support for our hypothesis.

*Chamas* leverages existing infrastructure to attempt to reach some of the most vulnerable members of Kenyan society – pregnant and postpartum women in poor, rural communities. The CHS provided an established workforce of trained CHVs who - with bolstered support, supervision, and mechanisms to increase work efficiency - significantly improved MNCH outcomes. Several CHV-related barriers to effective implementation of the current CHS model have been identified, including: the absence of consistent supervision, inadequate health training, poor linkages to health facilities, lack of accountability, and absence of remunerative pay.^28^ Our data support that by providing CHVs with additional oversight, a structured curriculum, and an opportunity to economize their time, the *Chamas* program helped narrow the margin between aspirational and achievable MNCH improvements. We anticipate that with added financial support, the *Chamas* program may yield even greater success. Moreover, we recognize opportunities for CHV financial remuneration are critical to ensuring the sustainability of this program and plan to prioritize this in future attempts to scale.

Our intervention approach – group-based women’s health education delivered by CHVs during pregnancy and postpartum – champions a theory of change that prioritizes three key areas: 1) empowering women with health and social literacy, 2) establishing a network of supportive peers, and 3) providing women with an opportunity to gain financial capital (*GISHE*). This third component is distinct from preceding strategies that promote a group-based, lay health worker-led model for MNCH.^12^ We suspect this multi-pronged approach that prioritizes these three critical components, in addition to leveraging an established CHV workforce, plays a significant role in enhancing positive outcomes. Evidence suggests peer support and peer accountability may enhance likelihood of practicing positive health behaviors.^29^ Further, there is a growing body of evidence that suggests coupling health education with microfinance may improve women’s health; however, most literature on associated reproductive health outcomes focus on contraceptive uptake and adherence to HIV/AIDS treatment.^30^ Among group members who participated in *GISHE*, we speculate the opportunity to generate savings likely served a dual purpose of motivating *Chamas* attendance while helping some participants overcome financial barriers to accessing care. Future analyses will attempt to dissect the influence of each of these components on overall program success. Lastly, while most programs intervene during distinct time periods (e.g. prenatal, intrapartum), *Chamas* embraces a life-course approach by engaging women throughout pregnancy and the first 1,000 days of their child’s life. We anticipate families who continue in *Chamas* likely experience health and social benefits throughout subsequent years. These effects are largely unexplored and will be the focus of future trials.

Preceding trials have examined the effect of similar community-based women’s health education groups on key MNCH behaviors, including facility-based delivery.^12^ A meta-analysis combining results from seven cluster randomized controlled trials conducted in resource-limited settings (i.e. Nepal, Bangladesh, Malawi, and India) found no evidence of intervention effects on facility-based delivery (OR 1.02, 95% CI 0.93, 1.12; I^2^ = 21.4%, 95% CI 0, 65.8%); similarly, these analyses revealed no evidence of effect on uptake of ANC or exclusive breastfeeding.^12^ Although data comparability is limited by differences in trial design, setting, and program structure (i.e. absence of table-banking), we observed significantly higher odds of facility-based delivery and exclusive breastfeeding in the intervention arm. Our findings strengthen evidence from an earlier *Chamas* pilot that similarly demonstrated increased odds of achieving these outcomes compared to the standard of care in rural western Kenya.^13^ Collectively, these findings highlight our intervention’s potential to improve MNCH outcomes by leveraging existing community health resources and infrastructure in settings like Kenya.

This work has several limitations. First, we experienced significant recruitment challenges that substantially reduced our sample size. Processes to contact eligible participants outside of health facilities proved arduous and complicated as locator data (e.g. home address, phone number) were often unreliable. This lost to follow-up between health facilities and communities likely introduced selection bias as we suspect women were not missing at random. It is possible these missing data reflect an inability to pay for cell phones (or data) or perhaps limited access in rural, hard-to-reach communities; thus, we may have failed to enroll some of the most socio-economically disadvantaged members of these communities. These challenges highlight a need to not only improve our strategy, but also to strengthen continuity between health facilities and communities to ensure vulnerable members of society are accounted for. Second, large amounts of missing data compromised the interpretability of certain outcomes, most notably infant immunizations. Despite established processes to monitor data quality, data collectors reported several challenges that compromised questionnaire completion. These obstacles included interruptions due to competing participant obligations (i.e. child care) and lack of a private interview setting. Further, relatively few participants possessed MOH Maternal Child Health booklets and among those who had them, few recorded data. This limitation may have introduced selection bias as mothers with completed records may have had greater access to or higher quality care. Alternatively, this could also reflect limited booklet availability, poor record-keeping, or other structural limitations worthy of further consideration. Finally, we observed a large amount of cluster heterogeneity, indicating significant, but anticipated community-level variation. Compositional effects within or between clusters such as proximity to health facilities, availability of service providers, and fidelity of program implementation may contribute to this variation. These effects may partly explain the unexpected attenuated difference in facility-based delivery in our sensitivity analysis of *Chamas* attendees. Continuing to clarify factors at the community level that contribute to variable outcomes may help bridge these observed geographic disparities.

These limitations are balanced by several noteworthy strengths of our study. As mentioned above, we detected significant results in our primary and secondary outcomes using an intention to treat approach. These observed effects were generally robust – e.g. not meaningfully changed following adjustment or imputation in our sensitivity analyses. The cluster randomized controlled design, implemented in a large and geographically diverse population, enhances generalizability of these findings. We saw no contamination across trial arms and minimized potential for information bias by masking data collectors, investigators, and analysts to cluster allocation throughout the trial. Further, by imposing relatively few exclusion criteria and a generous gestational age cut-off, we attempted to broaden inclusion to women who may have sought late ANC due to structural (e.g. distance to facility) or behavioral (e.g. delayed awareness of pregnancy) factors. Finally, it is worth noting that the proportion of facility-based deliveries, among other outcomes, was higher in both trial arms (80.9% intervention, 73.0% control) relative to county-level (58.3%) and national (61.2%) estimates.^1^ It is possible that study procedures, such as training and supervision, led to CHVs in control CUs being more likely to deliver standard of care, which might explain these observations.

*Chamas* offers an innovative approach to improve MNCH in resource-limited settings with significant health policy implications. This intervention demonstrated significant improvements in MNCH outcomes relative to the current standard of care; policy makers should take note of this strategy as they attempt to improve current initiatives. Since the program’s inception, we have emphasized the importance of collaboration with and investment from key stakeholders, including but not limited to: women, community leaders, CHVs, and MOH representatives at the county and national level. We respond to qualitative feedback from these stakeholders to ensure the program iteratively responds to the needs of its beneficiaries and remains community-driven. These commitments to collaboration and feedback inspire confidence in our program’s continued success. As we move towards scaling and integrating this intervention, our next steps will focus on addressing cost-effectiveness and enhancing adaptability to new settings.

In summary, *Chamas* participation significantly improved MNCH outcomes compared to the standard of care in western Kenya. This trial contributes robust data from sub-Saharan Africa that strengthens evidence to support community-based, women’s health education groups for MNCH in resource-limited settings.

## Data Availability

The de-identified data set and a data dictionary will be made available upon reasonable request with publication of the trial. Inquiries can be made to the corresponding author of this manuscript (Dr. Lauren Y. Maldonado).

## Contributions

ACD, LJR and JJS conceptualized, sought, and obtained funding for this study. LYM and MLS drafted the study protocol, developed data collection tools, and oversaw all data management processes led by GA. JB and LYM developed the statistical analysis plan, with critical feedback provided by all co-authors. AJ, JEI, and SC oversaw all research activities and coordinated research staff throughout the trial. JB conducted all statistical analyses with input from LYM. All authors assisted in interpreting results. LYM and JB authored the first draft of this article. All authors contributed to reviewing and editing the final draft of this article for important intellectual content. All authors approved submission of this manuscript for publication.

## Declaration of Interests

The authors of this manuscript declare no competing interests.

## Acknowledgements

We thank our study participants, community health volunteers, research assistants, and staff without whom this work would not be possible. We additionally thank the sub-county and county MOH representatives in Trans Nzoia for their support and collaboration. We are grateful for the mentorship and thoughtful feedback provided by our colleagues: Dr. Donald C. Cole (Dalla Lana School of Public Health, University of Toronto), Dr. K.S. Joseph (Department of Obstetrics and Gynecology, School of Population and Public Health, University of British Columbia), and Dr. Wendy Prudhomme O’Meara (Duke Global Health Institute, Duke University). This trial was made possible through the generous support of the Saving Lives at Birth (SL@B) partnership. The authors alone are responsible for the views expressed in this article and they do not necessarily represent the views, decisions, or policies of the SL@B partners or the institutions with which they are affiliated.

## Supplementary Files

*See separate attachment for supplementary files. Included attachments are listed below*.

- Supplementary Tables
- CONSORT Checklist (with extension for cluster randomized trials)
- Trial Protocol
- Statistical Analysis Plan

## Notes

### Competing Interest Statement

The authors have declared no competing interest.

### Clinical Trial

ClinicalTrials.gov (NCT03187873)

### Author Declarations

We received ethics approvals from the Institutional Research Ethics Committee at Moi University and Moi Teaching and Referral Hospital (IREC/2018/269) and Institutional Review Board at Indiana University (1905296355). We obtained written informed consent from all participants prior to data collection.

